# Growth Charts of Sutures and Fontanelles for The First Year of Infancy

**DOI:** 10.1101/2025.05.30.25328626

**Authors:** Siyuan Chen, Svein Kleiven, Ingemar Thiblin, Xiaogai Li

## Abstract

**AIM:** To establish normative growth charts for cranial sutures and fontanelles and to quantify the prevalence and morphology of accessory sutures during the first year of infancy.

**METHOD:** We retrospectively collected 194 head CT scans of infants (0–12 months) from the New Mexico Decedent Image Database. Using an automated morphometric framework (ISFMA), morphometric parameters such as suture width, length, sinuosity, and fontanelle area were measured. Growth trajectories were modelled using Generalized Additive Models for Location, Scale, and Shape (GAMLSS). Suture closure was quantified using the Suture Closure Ratio (SCR), and accessory sutures were identified and measured by type and age.

**RESULTS:** Suture length increased and width decreased with age. The metopic suture exhibited the earliest closure, while other sutures remained largely open throughout the first year. The anterior fontanelle area peaked around 3 months and gradually decreased thereafter. Accessory sutures, particularly the mendosal suture and superior median fissure, were common in neonates but diminished rapidly with age.

**INTERPRETATION:** This study provides the first comprehensive morphometric reference for infant cranial sutures and fontanelles. These normative data enhance clinical capacities for diagnosing craniosynostosis, evaluating abnormal skull development, and supporting forensic investigations involving abusive head trauma.

## Introduction

Cranial sutures and fontanelles are crucial anatomical structures of the human skull, intimately involved in cranial growth and brain development^1,2^, while also playing an important role in absorbing impact energy^3^. Craniosynostosis, characterized by premature suture fusion, commonly results in benign cranial malformation^4^, but if left untreated, it may lead to inhibited brain growth and long-term cognitive and psychological^5,6^. The evaluation of anterior fontanelle (AF) size in neonates has become a routine part of modern paediatric assessment^7^, as abnormalities in fontanelle morphology, especially the anterior fontanelle, can initiate in the early developmental stage, serious conditions such as achondroplasia, congenital hypothyroidism, and chromosomal defects like Down syndrome^8^.

Growth charts, widely used for monitoring childhood development, allow pediatricians, nurses, and parents to assess developmental delays or abnormalities^9^. The World Health Organization (WHO) and Centers for Disease Control (CDC) have provided growth charts for several individual growth characteristics such as head circumference and stature. Besides these general anthropometric traits, some research focusing on specific anthropometric variable, such as global brain phenotypes, providing valuable tools for assessing brain development^10^, yet no growth chart on cranial suture and fontanelle development. A number of previous studies have examined infant cranial sutures and fontanelles using medical imaging techniques^11–17^. However, most of these investigations rely on inconsistent measurement focusing on isolated features at one specific age, such as suture width, fusion timing, or the fontanelle size. To date, no study has systematically established comprehensive sutures and fontanelles growth charts across the first year of infancy. This gap not only limits pediatricians’ understanding of normal development of sutures and fontanelles, but also hinders early identification of abnormalities, sometimes leading to unnecessary clinical interventions such as misinterpreting normal early anterior fontanel closure in normocephalic infants^18^.

Beyond general developmental patterns, structural anomalies such as accessory sutures present further challenges for diagnosis and forensic interpretation. Accessory sutures are rare developmental anomalies that commonly appear in the parietal and occipital bones, probably due to multiple ossification centers^19^. Although accessory usually asymptomatic^20^, recent biomechanics study has demonstrated that the presence of sutures may significantly influence cranial fracture patterns by absorbing impact energy and facilitating the initiation or propagation of linear fractures^21^. In some cases, accessory sutures can be difficult to distinguish from skull fractures, posing significant challenges for forensic identification. Current differentiation largely replies on qualitative radiological features, such as bilateral symmetry, widening along the suture, and correlation with clinical findings^19^. However, radiologists, pediatricians, and forensics lack of prevalence of different accessory sutures across pediatric age groups, let alone quantitative age-specific morphometrics such as accessory suture length.

The present study aims to establish a comprehensive normative reference charts for suture and fontanelle during the first year of infancy. We first conducted systematically consistent measurements of key morphometric parameters, including suture width, length, fontanelle area and other major cranial sutures and fontanelles characteristics. We then created suture and fontanelle charts using generalized additive models for location, scale and shape (GAMLSS), a robust and flexible method recommended by the World Health Organization (WHO) for modelling non-linear growth trajectories^22^. In addition, we also applied principal component analysis (PCA) to evaluate shape variation across age groups. Finally, we evaluated the prevalence and average length of different accessory sutures in the parietal and occipital bones across different infant age groups, aiming to establish reference charts to support both clinical and forensic assessment.

## METHOD

### Study design and CT data

We conducted a retrospective review of CT images from the New Mexico Decedent Image Database (NMDID)^23^. Subjects with conditions that could affect the normal suture development, such as prematurity, intrauterine fetal demise, maternal or fetal birth complications, and congenital anomalies including Down syndrome, were excluded. After screening, a total of 194 head CT scans from infants aged 0 to 12 months (106 males and 88 females) were included in the analysis.

### Anthropometric measurement of sutures and fontanelles

Morphometric analysis was conducted using the Infant Suture & Fontanelle Morphology Analysis (ISFMA) framework^24^, which enables automated extraction of parametric features from 800 semilandmarks identified along the borders of sutures and fontanelles on CT images (Figure 1). Suture length was defined as the linear distance between the anatomical endpoints of each suture. Suture sinuosity index was calculated by dividing the suture length by the straight-line distance between the endpoints. Suture width was calculated as the mean of the shortest distances between opposing borders at each semilandmark along a given cranial suture. Furthermore, fontanelles surfaces were reconstructed using a thin-plate-spline interpolant, enabling calculation of surface area and curvature. A total of 28 morphometric parameters were measured (Table 1), with technical details described in our previous study^24^.

**Table 1.**
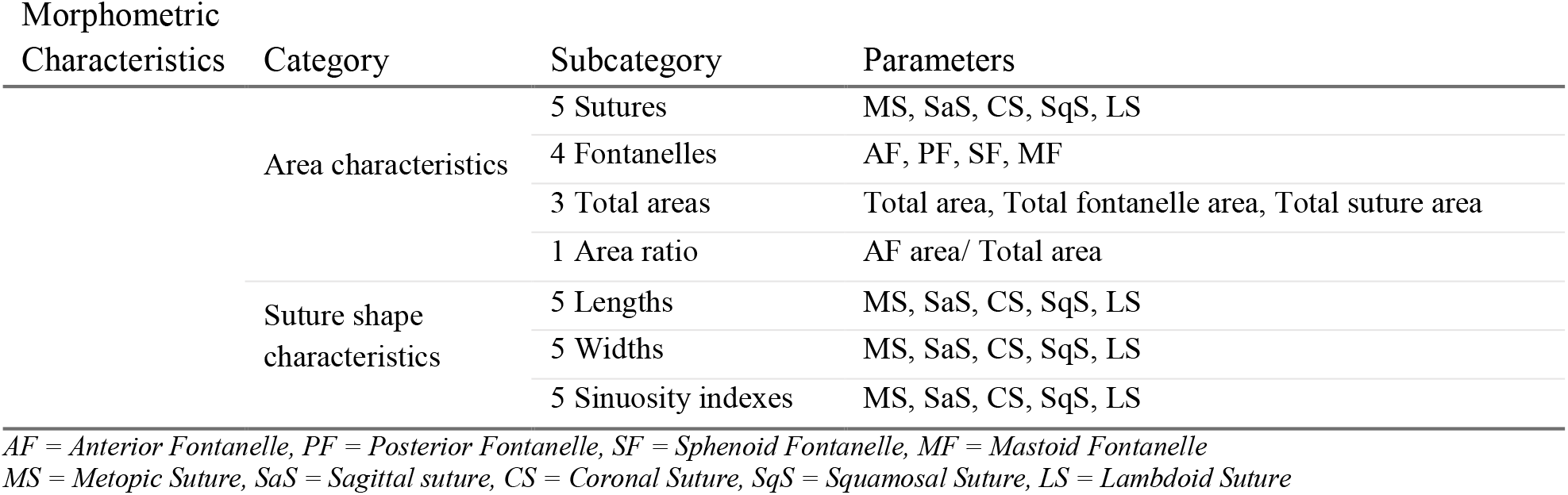
28 morphometric characteristic measured in this study.

| Morphometric Characteristics | Category | Subcategory | Parameters |
| --- | --- | --- | --- |
|  | Area characteristics | 5 Sutures | MS, SaS, CS, SqS, LS |
|  |  | 4 Fontanelles | AF, PF, SF, MF |
|  |  | 3 Total areas | Total area, Total fontanelle area, Total suture area |
|  |  | 1 Area ratio | AF area/ Total area |
|  | Suture shape characteristics | 5 Lengths | MS, SaS, CS, SqS, LS |
|  |  | 5 Widths | MS, SaS, CS, SqS, LS |
|  |  | 5 Sinuosity indexes | MS, SaS, CS, SqS, LS |

AF = Anterior Fontanelle, PF = Posterior Fontanelle, SF = Sphenoid Fontanelle, MF = Mastoid Fontanelle
MS = Metopic Suture, SaS = Sagittal suture, CS = Coronal Suture, SqS = Squamosal Suture, LS = Lambdoid Suture

**Figure 1.**
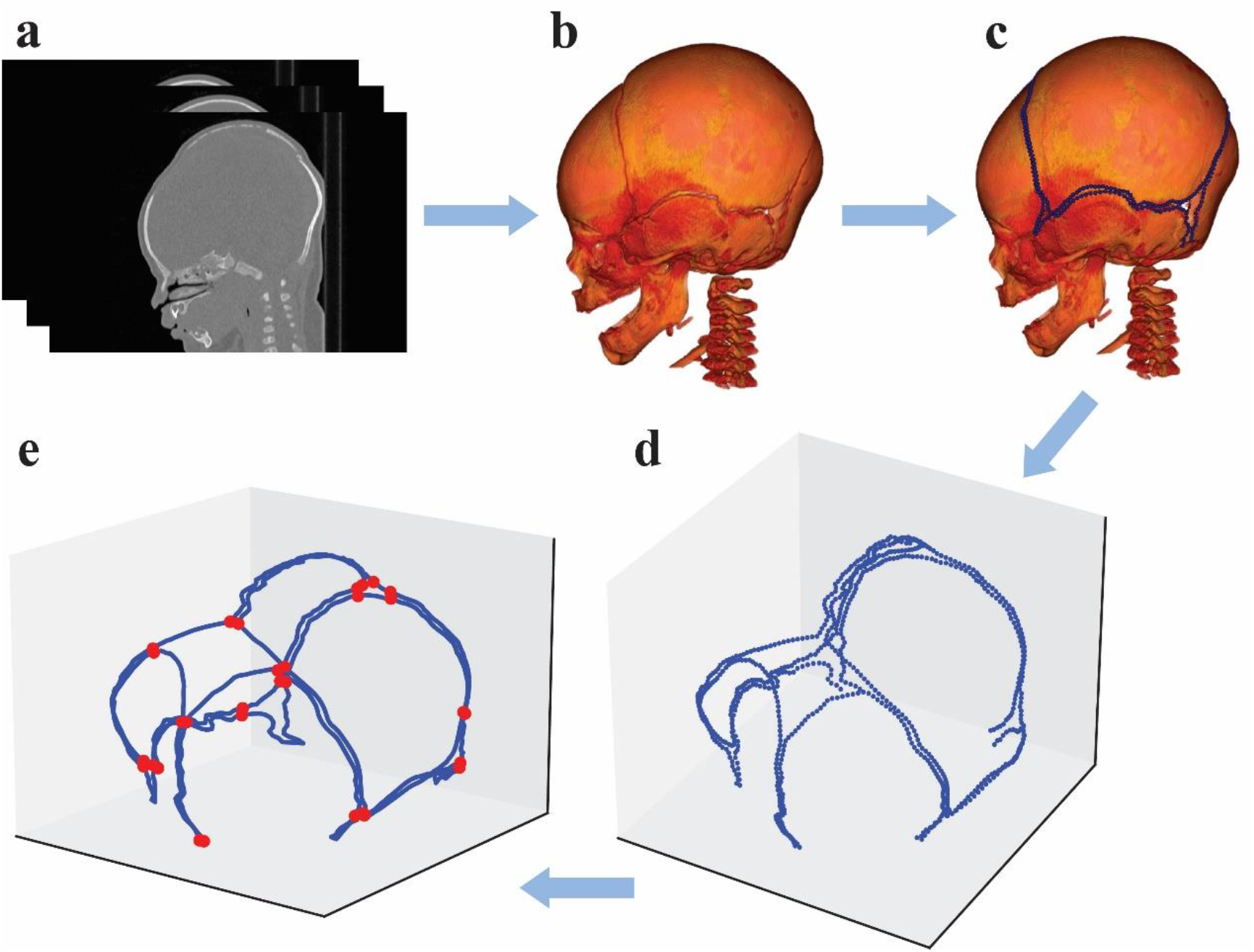
Procedure of identifying semilandmarks and junction points from CT images. a Acquisition geometry regularization and head posture alignment of CT image. b Segmented skull. c Semilandmarks identification along the borders of infant body plate bones (marked as blue dots). d In total 800 semilandmarks were used to calculate morphological traits of sutures and fontanelles. e 32 junction points (marked as red dots) were identified from the 800 semilandmarks to determine the borders of cranial sutures and fontanelles according to the width variances.

### Suture closure analysis

Cranial sutures are typically patent at birth and gradually narrow over time. Based on the visual resolution threshold of the human eye (approximately 0.2 mm)^25^, we used 0.1 mm as a practical threshold for suture closure on CT images. Widths below this threshold were defined as radiologically “closed”, though histologically they may still be patent. Thus this threadhold reflects image-based closure, not histological fusion. Suture closure ratio (SCR) was defined as the percentage of the closed length over total suture length. Madeline and Elster^26^ proposed a five-grade system for assessing infant suture closure based on visual inspection of CT images, we adapted this grading framework to a quantitative classification scheme based on SCR. This approach is illustrated using the metopic suture as an example (Figure 2).

**Figure 2.**
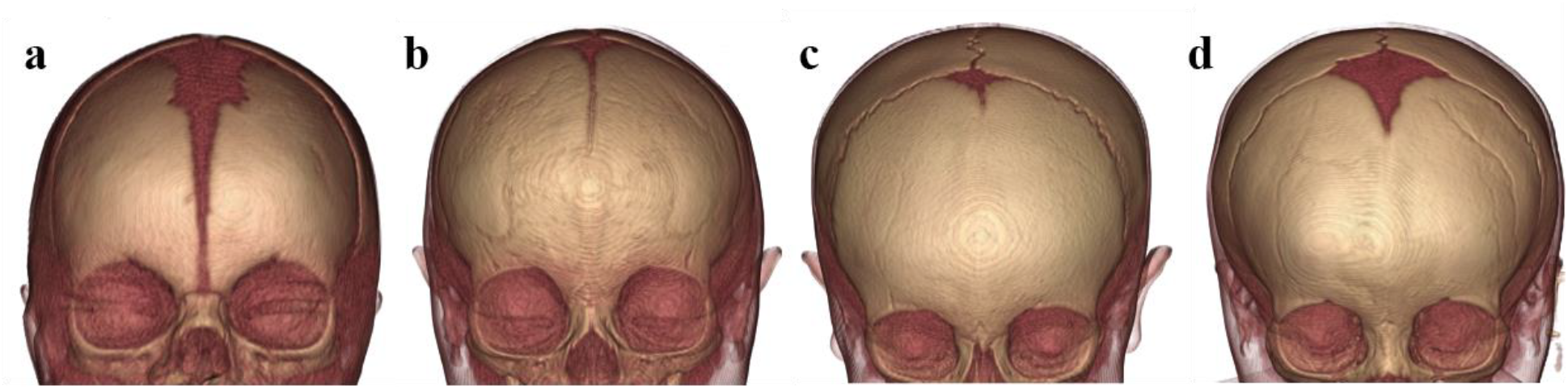
Suture closure ratio grading system in the metopic suture, where Grade 1 = no closure along the whole length with SCR = 0%; Grade 2 = partial closure with 0% < SCR ≤10%; Grade 3 = moderate closure with 10% < SCR < 90%; Grade 4 = substantial closure with 90% ≤ SCR < 100%; Grade 5 = complete closure with SCR = 100%. The figure for Grade 2 is missing due to the unavailability of a corresponding subject. **a** Subject of Grade 1 with SCR = 0%. **b** Subject of Grade 3 with SCR = 55%. **c** Subject of Grade 4 with SCR = 91%. **d** Subject of Grade 5 with SCR = 100%.

### Accessory suture measurement

Lengths and prevalence of five accessory sutures were measured: the mendosal suture (MS), superior median fissure (SMF), parietal transverse accessory suture (PTAS), parietal accessory suture near the lambdoidal suture (PASLS), and parietal accessory suture near the squamosal suture (PASSS). Age-specific prevalence across different ages and mean lengths were quantified from CT images.

### Normative growth trajectories of sutures and fontanelles

Normative growth trajectories were generated for major cranial sutures and fontanelles by sexes, including centile values for suture length, width and fontanelle areas, using GAMLSS. GAMLSS is an extension of generalized linear model (GLM) that allows for modelling multiple parameters as functions of explanatory variables. This method is well suited for capturing non-linear growth trajectories and is recommended by the WHO for constructing biological growth standards.

### Shape variation and regression analysis

Prior to PCA, generalized procrustes analysis (GPA) was applied to the geometric coordinates of 800 semilandmarks to rigidly remove the influence of incomparable spatial positions without altering their size, followed by a sliding thin-plate spline (TPS) procedure to establish geometric homology of the 800 semilandmarks across subjects. The *Morpho* package in the statistical software R was subsequently used to slide these aligned semilandmarks along each target borderline using the sliding TPS algorithm. Finally, multivariate regression analysis was performed to construct a statistical model incorporating subject-level covariates, including age, suture width, suture length, and fontanelle area, to visualize morphological variation in sutures and fontanelles across percentiles and age groups.

## RESULTS

### Growth charts of sutures and fontanelles

Coordinates of semilandmarks for all subjects are documented in Supplementary Material A. Figure 3 illustrates the normative growth trajectories and centile curves for suture length and width, as well as the proportions of corresponding suture closure grades at different ages based on corresponding SCR values. Across all suture types, a consistent developmental trend is observed: length increases while width decreases during the first year of life. Figure 4 presents the normative trajectories of fontanelle area. AF) displayed a distinct pattern to other fontanelles, reaching a maximum area of 745.02 mm^2^ around 3 months of age, while other fontanelles gradually close with age. Additional morphological results and detailed growth charts are provided in Supplementary Material B and C.

**Figure 3.**
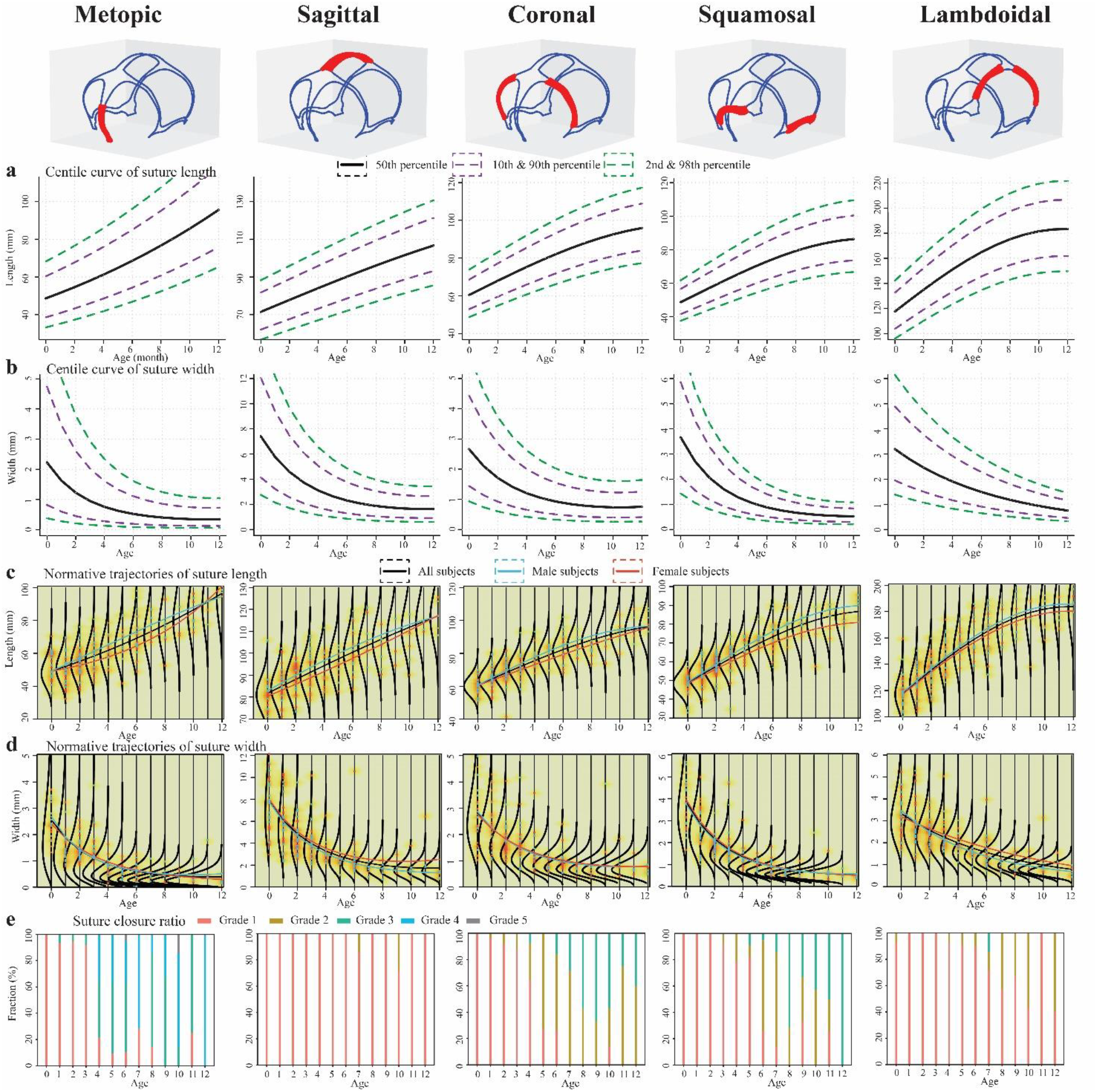
a Different centile curves for suture length during the first year of infancy. The median curve is shown in black, with the 10% and 90% centiles denoted by purple dotted lines, and the 2% and 98% centiles by green dotted lines. b Different centile curves for suture width during the first year of infancy. c Normative suture length trajectories during the first year of infancy. The density of the data points is represented by the color chromaticity. The male data are plotted in blue, and the female data are plotted in red. Three center lines in black, blue and red are respectively fitted by GAMLSS method using all data, only male data, and only female data. The other curves are probability density distributions for each age group. d Normative suture width trajectories during the first year of infancy. e Fraction of suture closure ratio grades of various cranial sutures in the first year, where Grade 1 means totally open and Grade 5 means completely fused.

**Figure 4.**
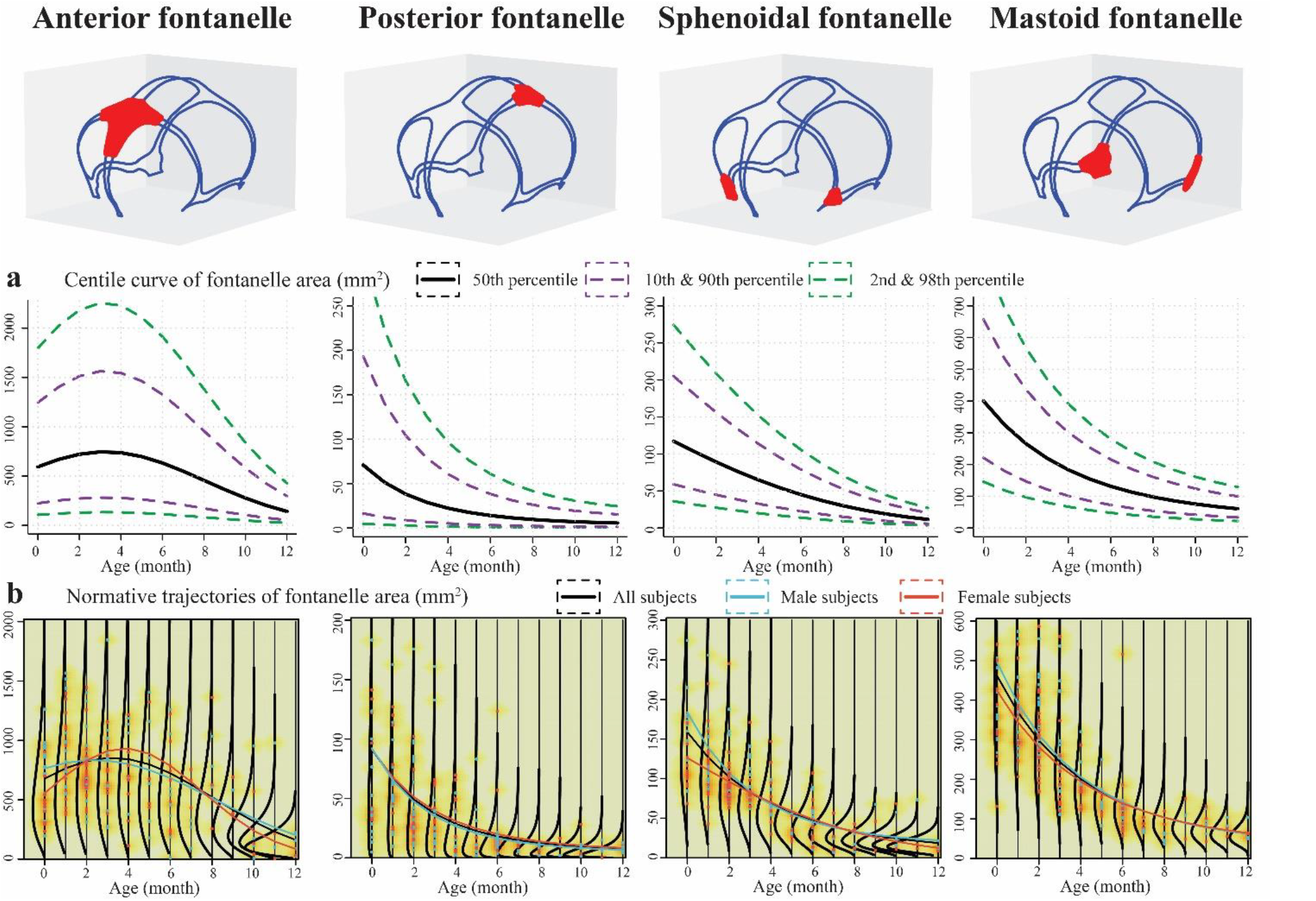
a Different centile curves for fontanelle area. b Normative fontanelle area trajectories of infant in different age.

### Morphological variation across percentiles

The first 35 principal components (PCs) captured over 95% of variance in suture size, closure, sinuosity, and fontanelle shape, reducing dimensionality from 2400 to 35 features. By assigning percentile-based covariates values for each age group, Figure 5 illustrates the morphological differences between subjects at the 10th and 90th percentiles across infancy.

**Figure 5.**
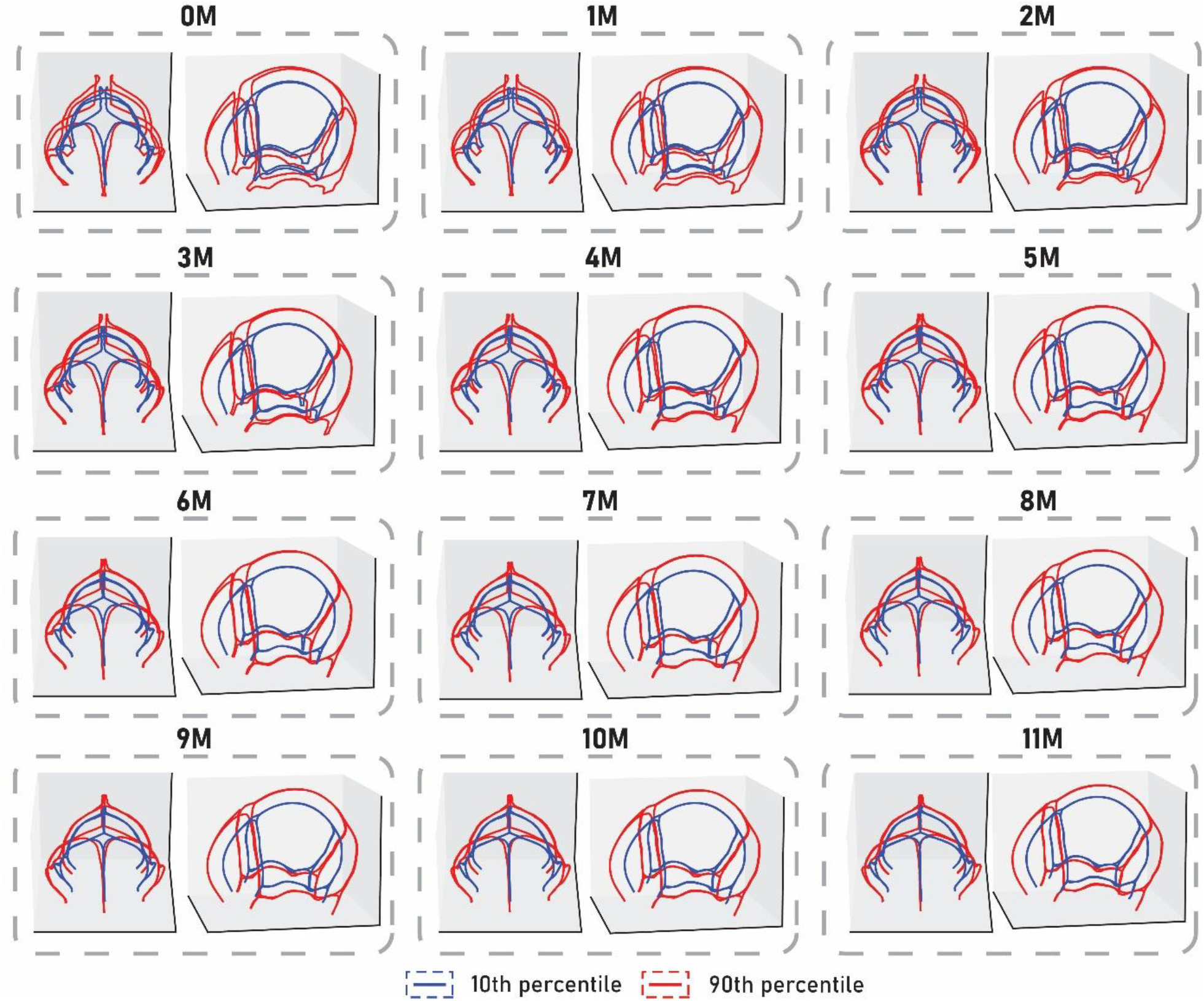
Morphological variations across different age groups of infants.

### Accessory sutures: prevalence and variation

We identified five types of accessory sutures, including two occipital types (MS, SMF) and three parietal types (PTAS, PASLS, PASSS) (Figure 6).

**Figure 6.**
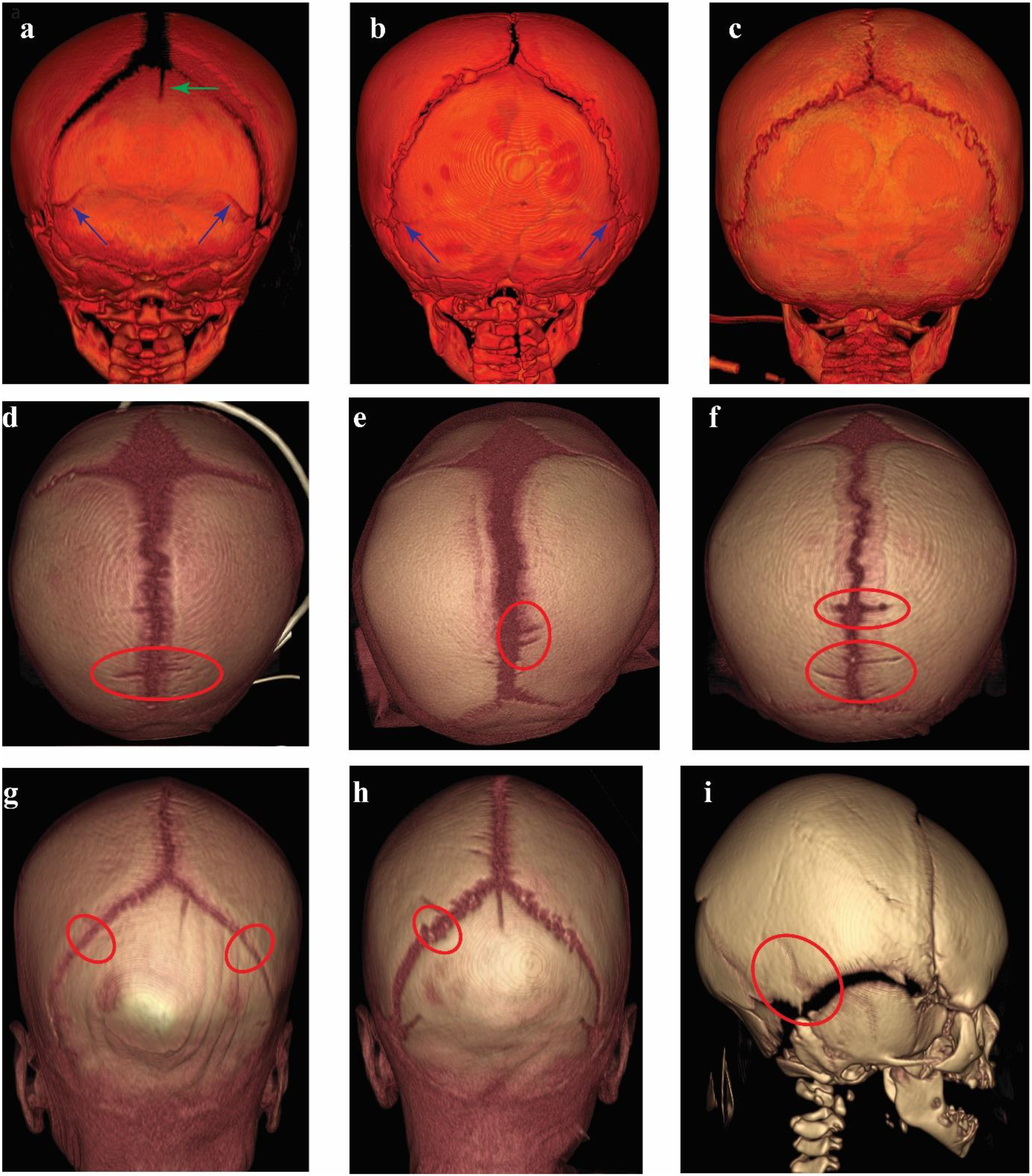
3D-CT reconstruction images of the skulls with different types of accessory sutures. a A newborn with a superior median fissure (green arrow) and mendosal sutures (blue arrows). b A 4MO infant with only mendosal sutures (blue arrows). c An 8MO infant without a superior median fissure and mendosal suture. d Bilateral parietal transverse accessory suture. e Unilateral parietal transverse accessory suture (PTAS). f Multiple parietal transverse accessory suture. g Bilateral parietal accessory suture near lambdoidal suture (PASLS). h Unilateral parietal accessory suture near lambdoidal suture. i Parietal accessory suture near squamosal suture (PASSS).

Table 2 demonstrates their frequency and length by age. SMF was no longer observed beyond 4 months, and MS disappeared after 7 months. Among 0-month-old infants, the frequency of MS and SMF was 100% and 89%, respectively. Compared with occipital accessory sutures, parietal accessory sutures were considerably less common. Notably, PASSS was the rarest, identified in only one of the 194 subjects (0.5%).

**Table 2.** Relationships between the age and frequency (F) and length of different types of occipital accessory sutures and intraparietal accessory sutures.^1^.

| Age (month) | Occipital accessory sutures |  |  |  | Intraparietal accessory sutures |  |  |  |  |  |
| --- | --- | --- | --- | --- | --- | --- | --- | --- | --- | --- |
|  | MS |  | SMF |  | PTAS |  | PASLS |  | PASSS |  |
|  | F | AL | F | AL | F | AL | F | AL | F | AL |
| 0 | 100 | 12.4 | 88.9 | 8.7 | 48.1 | 6.3 | 18.5 | 6.8 | 3.7 | 7.2 |
| 1 | 100 | 11.7 | 70 | 7 | 33.3 | 6.4 | 3.3 | 4 | 0 | 0 |
| 2 | 83.3 | 10.3 | 41.7 | 6.7 | 11.1 | 5.2 | 2.8 | 5.5 | 0 | 0 |
| 3 | 54.2 | 12.4 | 20.8 | 6.1 | 16.7 | 4.6 | 8.3 | 6 | 0 | 0 |
| 4 | 42.9 | 11.7 | 0 | 0 | 0 | 0 | 7.1 | 19.4 | 0 | 0 |
| 5 | 36.4 | 7.3 | 0 | 0 | 0 | 0 | 0 | 0 | 0 | 0 |
| 6 | 31.6 | 6.8 | 0 | 0 | 0 | 0 | 0 | 0 | 0 | 0 |
| 7 | 42.9 | 3.9 | 0 | 0 | 0 | 0 | 0 | 0 | 0 | 0 |
| over 7 | 0 | 0 | 0 | 0 | 0 | 0 | 0 | 0 | 0 | 0 |

<sup>1</sup>(F: Frequency (%). AL: Average length of subjects with corresponding accessory sutures (mm). The length of the accessory sutures was measured as the distance between two endpoints in the CT image. MS: Mendosal suture. SMF: superior median fissure. PTAS: Parietal transverse accessory suture. PASLS: parietal accessory suture near lambdoidal suture. PASSS: Parietal accessory suture near squamosal suture.)

## DISCUSSION

This study presents the first normative growth charts for infant cranial suture and fontanelle development during the first year of life using a consistent, automated morphometric approach. We also quantified the frequency and length variation of accessory sutures across age groups. Normative growth trajectories of major cranial sutures and fontanelles were developed for both sexes, revealing substantial individual variability in morphology and clear differences in growth and closure rates across sutures and fontanelles. These findings, together with our accessory suture analysis, provide a reference framework for more accurate clinical assessments of developmental anomalies and offer a foundation for future work expanding normative datasets, exploring sex-specific developmental trajectories.

### Consistency and discrepancies with other studies

Although few in number, previous studies have reported certain suture types or AF size in infants. To our knowledge, this study is the first to offer comprehensive morphometric measurements for all major cranial sutures and fontanelles during the first year of infancy. A comparison between some of our results and previous reports is presented in Table 3.

**Table 3.**
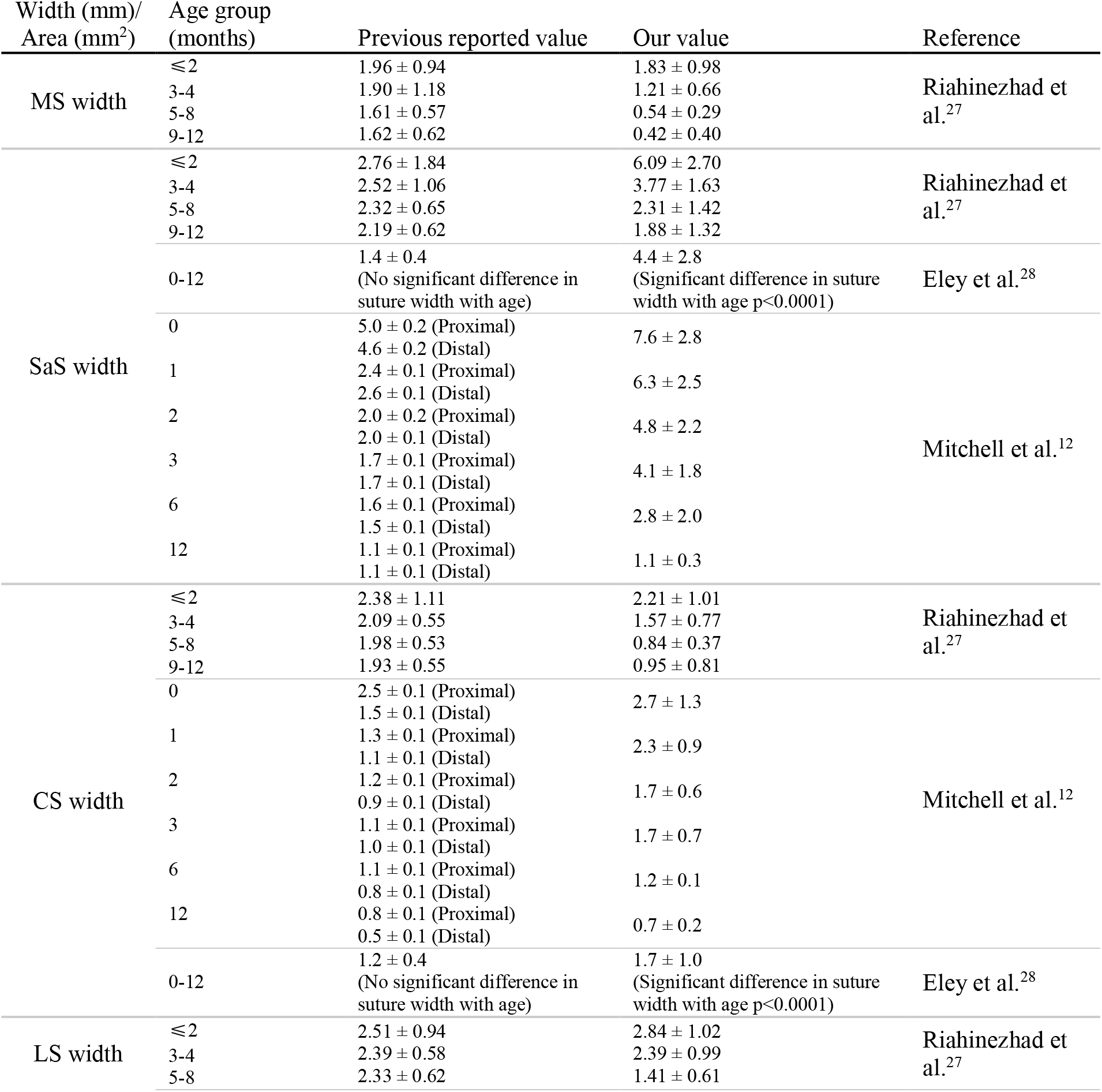
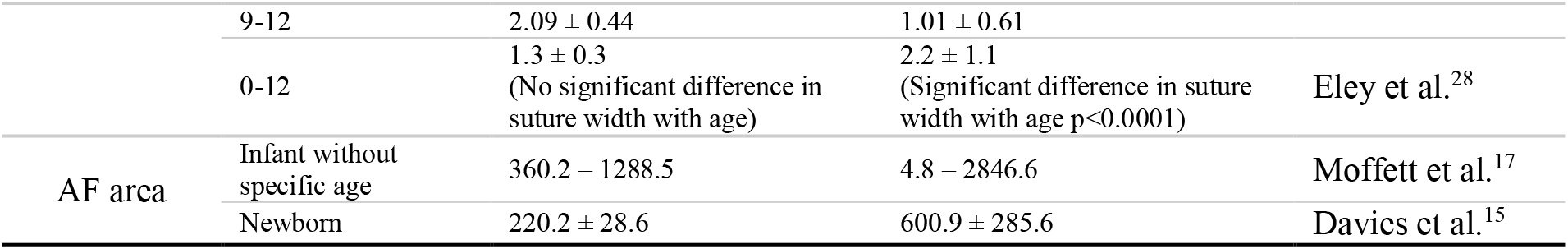
Comparison of Cranial Suture Widths and Anterior Fontanelle Area in Infants between this study and previous reports.

Apart from the sagittal suture, our measurements generally align with previous findings in younger infants but diverge significantly in older age groups. For example, when compared with Riahinezhad et al.^27^, our measurements showed greater age sensitivity. These differences may stem from ethic variation and methodological factors. Previous studies often used CT slice-based width estimation at selected locations, a process prone to introducing observer bias. In contrast, our method defined suture boarders automatically and computed average width across the full suture length, enabling consistent measurements and reducing subjective error. As the widest suture, the sagittal suture typically remains open through 12 months. The similarity of the sagittal suture width at older ages supports this as wider sutures may partially mitigate the marking errors introduced by visual estimation. Radiologists should exercise more caution when evaluating infant sutures across different types of imaging modalities. Contrary to common understanding and our findings, measurements of suture width based on MRI images suggest no significant difference in suture width with age, which is highly unreliable due to the poorly defined boundaries of cranial sutures on MRI.

In line with previous reviews of suture closure patterns, the metopic suture, as the earliest cranial suture to close, typically undergoes fusion between 3 and 11 months of age, while the other major cranial sutures generally remain unfused throughout the entire infancy^1^. Our findings support this timeline, with over 90% of individuals demonstrating signs of metopic closure beginning in the fourth month, and complete fusion observed in some cases as early as six months. All subjects exhibited closure ratios exceeding 90% by 12 months. No other cranial sutures in our cohort reached a SCR grade of 4 or higher during the first year. The sagittal suture, in particular, showed initial signs of closure only after seven months of age, and most individuals retained a fully open sagittal suture throughout infancy, with maximum SCR values remaining below 10%. These observations reinforce the distinct closure trajectory of the metopic suture and highlight the prolonged patency of the other major cranial sutures in early postnatal development.

### Anterior fontanelle area and measurement

Comparison of AF area highlights the importance of standardized measurement approaches. Traditional methods estimate fontanelle area using longitudinal and transverse diameters derived from four fontanel “corners”, assuming a quadrilateral shape^15,16^. Moffett et al.^17^ proposed a 3D reconstruction method followed by the manual selection of voxels corresponding to the AF in the CT image. However, ambiguity in suture and fontanelle boundaries remain a challenge.

Our study identified AF borders using consistent geometric rules, and subsequently the surface area was calculated based on 3D reconstructions. Interestingly, although the peak age of AF size reported in our study (between 2 and3 months) differs from that reported by Duc and Largo^16^ (peak between 1 and 2 months), their research also revealed an initial enlargement of AF size during early infancy. Though the ossification of infant sutures continues throughout the developmental period, we suggest the rapid increase in head circumference during early infancy is the primary contributor to the increase of the absolute AF area. As head circumference growth decelerates, the closure of the AF becomes dominant, resulting in a gradual decrease in its area thereafter. Other fontanelles dit not exhibit this trajectory.

### Accessory suture

Accessory sutures must be considered when evaluating cranial morphology, especially in neonates. Most newborns in our sample exhibited at least one accessory suture. Our findings align with Muron et al^29^, who reported that MS and SMF typically occur before 5 months, with median presence at 1 and 0 months, respectively. Our charts complements the key length information across different ages.

Ignoring accessory sutures can mislead both clinical and forensic investigations, particularly in suspected cases of abusive head trauma, where accessory sutures and linear fractures^19,20^ can resemble linear skull fractures due to the similarity in radiological images. Moreover, this study highlights the need for additional biomechanicscs research on accessory sutures. Existing infant head models, such as finite element head models, do not explicitly account for accessory sutures^30^, which may affect the conclusions drawn from numerical simulations, especially for newborns. One of the next steps would be to conduct comprehensive biomechanics studies on this topic, as the required quantitative anatomy information has been provided by this study.

## Limitations

The first limitation of our study is the sample size and demographic diversity. NMDID only includes CT scans of individuals who died between 2010-2017 in New Mexico, with white being the dominant race. The 12-month age limit was selected in this study because, beyond this age, the number of available CT scans was too limited to support meaningful statistical modeling. Thus, our analysis focused on the first year, which is a critical window for cranial suture and fontanelle development. Although we found differences in growth trajectories between males and females, a larger sample size with balanced data across genders is needed to better assess significant differences in growth charts for different genders. In addition, we were unable to capture true biological suture closure. A suture width of 0.1 mm does not necessarily represent complete biological fusion. However, given that the spatial resolution of CT imaging is far lower than the microscopic scale of actual biological fusion, a 0.1 mm-wide suture in CT reconstructions is considered fully ossified (Figure 2). Despite these limitations, this study exploited all available sample data and provided new and valuable insights into infant suture and fontanelle growth, as well as accessory suture development during the first year of infancy.

## CONCLUSION

To our knowledge, this is the first study to comprehensively chart cranial suture and fontanelle development throughout the first year of infancy. The resulting normative growth charts and quantitative analysis of accessory sutures provide practical tools for clinical diagnostic, radiologic evaluation and forensic investigation. These findings also lay essential groundwork for expanding normative anatomical references and integrating accessory suture data into biomechanical modeling.

## Data Availability

All data produced in the present work are contained in the manuscript.

## Notes

### Competing Interest Statement

The authors have declared no competing interest.

### Funding Statement

This research has received funding from the Swedish Research Council (VR-2016-04203, VR-2020-04496 and VR-2020-04724).

### Author Declarations

The Institutional Review Board of the University of New Mexico gave ethical approval for the creation and research use of the New Mexico Decedent Image Database (NMDID). Access to the database was granted upon institutional request. All data used in this study were fully de-identified and derived from deceased individuals. No additional ethical approval was required for this secondary analysis.

